# Predictors of long-term progression to chronic kidney disease in HIV infection in Ghana from 2003-2018

**DOI:** 10.1101/2022.11.23.22282665

**Authors:** David R Chadwick, Fred Barker, Colette Smith, Okyere Perditer, Yasmine Hardy, Dorcas Owusu, Giovanni Villa, Fred Stephen Sarfo, Anna-Maria Geretti, Richard Phillips

## Abstract

**Aim:** HIV is associated with an increased risk of progression to chronic kidney disease (CKD), and this risk is higher in people of West African descent than many other ethnicities. Our study aimed to assess the rates of progression to CKD and predictors of rapid progression in patients receiving antiretroviral therapy (ART) in central Ghana between 2003 and 2018.

**Methods:** This single-centre retrospective study enrolled people with HIV (PWH) initiating ART in Ghana between 2003-2018. Demographics, hepatitis B (HBsAg) status, ART regimens and eGFR measurements were recorded, and multi-level model linear regression was performed to determine predictors of greater levels of eGFR decline.

**Results:** 659 participants were included in the study with a median follow-up time of 6 years (IQR 3.6-8.9). 149 participants (22.6%) also had confirmed HBV co-infection. Tenofovir was associated with the highest mean rate of eGFR decline of all Nucleoside/Nucleotide Reverse Transcriptase Inhibitors (NRTIs), representing a statistically significant annual decline -1.08 mL/min/1.73m^2^/year (CI: -0.24, -1.92) faster than those taking zidovudine. Regarding other ARTs, both nevirapine (-0.78mL /min/173m^2^/year; CI: -0.17, -1.39) and protease inhibitors (-1.55mL/mil/173m^2^/year; CI: - 0.41, -2.68) were associated with slower eGFR declines compared with efavirenz. Negative HbsAg status was associated with greater eGFR decline compared with positive HBsAg status (-1.25mL/mil/173m^2^; CI 0.29. 2.20).

**Conclusion:** Increased rates of eGFR decline amongst PWH in Ghana were associated with tenofovir, nevirapine, and protease inhibitor use as well as negative HBsAg status. Further higher-quality research is needed to explore long-term predictors of eGFR decline in African populations.

## Introduction

People with HIV (PWH) are at increased risk of chronic kidney disease (CKD) secondary to disorders such as HIV-associated nephropathy (HIVAN), HIV immune complex glomerulonephritis, diabetes and hypertension. Additionally, antiretroviral treatments (ART) for HIV have also been shown to adversely affect renal function in PWH^1-9^, though on balance the use of ART has considerably decreased both HIV-associated CKD and progression to end stage renal disease (ESRD)^10-13^. However, renal disease remains common amongst PWH and those affected are at higher mortality risk than those with good renal function^,15^.

Africa has the highest prevalence of HIV-associated CKD (estimated glomerular filtration rate [eGFR] < 60 ml/min) globally with West Africa being the most affected region (15%) compared to Southern Africa (3%)^16^. Amongst PWH of African ethnicity in the UK, West Africans have also been found to be at highest risk of ESRD^17^. On the African continent, major factors contributing to CKD progression include genetic predisposition^18-21^ and other co-infections such as hepatitis B virus (HBV)^22^, which affects approximately 14%-17% of PWH in Ghana^23,24^. Studies have noted that chronic HBV infection is associated with CKD or progression to CKD both in PWH^22,25^ and other populations^26^. However, there remains a paucity of long-term research exploring this relationship in African co-infected populations with only one study evaluating it specifically as a possible risk factor for CKD progression^27^.

The nucleotide reverse transcriptase inhibitor tenofovir disoproxil fumarate (TDF) is active against both HIV and HBV. It has been frequently and widely used in Sub-Saharan Africa as a first-line ART for over a decade. Despite modest evidence of improved side effect profiles over some other common antiretrovirals^28,29^, the drug has demonstrated associations with impaired glomerular filtration and tubular dysfunction (including Fanconi’s syndrome) in HIV positive and negative populations^30-34^. A systematic review of African studies showed statistically significant positive relationships between TDF use and renal dysfunction in around a quarter of its included studies^35^. However, few published studies have assessed how renal function changes over long periods of time; only three studies to date assessed participants on ART for a duration of >5 years ^36-38^. One of these studies noted a significantly greater eGFR decline in the TDF group compared with the control^37^.

Our single-centre study was designed to determine risk factors associated with developing CKD or fast eGRF decline in Ghanaian PWH taking ART for up to 15 years. Our specific focus was the influence of factors such as individual ART drugs, including TDF exposure, and HBV co-infection on renal function decline.

## Patients and Methods

### Settings

The study was conducted at the Komfo Anokye Teaching Hospital (KATH) in Kumasi, Ghana. The Committee on Human Research Publications and Ethics at Kwame Nkrumah University of Science and Technology (KNUST) approved this study. All PWH provided written consent to take part in the study.

### Study population and therapy

The majority of participants were enrolled from previous studies with retrospective data collection starting in 2004 and completed in 2019^14,27,39^, alongside a small convenience sample of PWH attending the clinic in February 2019 who were enrolled explicitly for this study. The study included adults (18 years or older) who had both a basal creatinine measurement (within 3 months of starting ART) and between one and three further result beyond this date.

National HIV treatment guidelines (2003) originally recommended that PWH were initiated on either zidovudine or stavudine with lamivudine, plus either nevirapine or efavirenz, with some protease inhibitors, didanosine and abacavir available for second-line therapy. Once TDF became available in 2010 a small proportion of patients, including most of those identified with hepatitis B co-infection, were commenced on or switched to TDF.

### Data collection and definitions

All demographic, clinical and laboratory data used were collected from clinic case notes or research registers. Participants’ first recorded ART regimens were used to define their ‘baseline’ regimen. Data were recorded according to the type of ART used and the length of time it was taken for. Cases where a patient switched between different ART medications were recorded for later data analysis.

The creatinine data used in this study was mostly from samples as part of routine or standard hospital care (locally measured), or from one of several research studies (measured in the UK). Routine management of PWH in KATH did not include regular blood creatinine measures, rather these were done periodically or when felt to be clinically indicated. Likewise, until recently most PWH were not routinely tested for hepatitis B surface antigen (HBsAg) unless they were found to have deranged transaminase measurements. None-the-less a significant number of patients attending the clinic were tested for HBsAg and had more frequent creatinine measurements as part of previous studies. HIV viral load is also not routinely measured and was not included in any analyses.

PWH were considered to be hepatitis B co-infected if they had at least one positive HBsAg test recorded. eGFR was calculated using the CKD-EPI creatinine equation with standard adjustment for gender and black race^40^. In line with previous analyses of HIV-positive populations^41^, rapid eGFR progression was defined as a change in eGFR of greater than 5 ml/min/1.73m^2^ per year between baseline and final eGFR measurement.

### Statistical analysis

Two study populations were considered for analysis: (i) the sub-set of 545 participants with at least three years’ follow-up, chosen to ensure medium-to-long-term eGFR changes were being evaluated; (ii) all 659 participants with at least two eGFR measurements for formal regression analysis, to maximise the use of available data as well as to minimise risk of bias associated with loss-to-follow-up. Among the 545 participants with at least 3 years’ follow-up, the annualised eGFR change (the difference between the first and last eGFR measurements, divided by the length of follow-up in years) was calculated and compared according to antiretroviral use and hepatitis B (HBsAg) status using analysis of variance (ANOVA). In addition, the percentage that had experienced rapid progression over >3 years’ follow-up, as defined above, was calculated stratified by hepatitis B and ART status and compared using a chi-squared test. Next, considering all 659 participants, a multi-level model was constructed to investigate factors associated with eGFR changes over time. Time was considered as a linear association. Although covariates were also included as main effects, the results focus on the association between covariates and the rate of eGFR change over time (that is the covariate-time interaction terms). An unstructured correlation matrix was used, and random effects terms were included for the intercept were performed using SAS Version 9.4 (SAS Institute Inc, Cary, NC).

### Sensitivity analysis

Sensitivity analysis assessed whether results remained consistent in a restricted cohort where only participants with at least three years between their first and last eGFR result were included.

## Results

### Study population characteristics

After review of research databases (n=463) and the convenience sample (n=196), 659 PWH on ART were identified with two or more eGFR measurements available, of which 545 had at least 3 years follow-up between first and final measure. The characteristics of this population are described in Table 1. The median time between first and final creatinine measurement was 6 years (IQR 3.6-8.9). 149 participants (22.6%) had confirmed HBV co-infection (HBsAg positive), 210 (31.9%) were HBsAg-negative and the remainder had unknown HBV status. 77 PWH (11.7%) were initiated on TDF-based ART with the majority of patients starting zidovudine-based ART (59.3%). In terms of typical ‘third’ ARTs, efavirenz was the most commonly used drug initially (59.3%) followed by nevirapine (35.8%) and protease inhibitors (4.9%). At baseline, the mean eGFR was 104.5 ml/min/1.73m^2^ (SD 31.4; range 11.9 to 185.2). 55 (8.4% of) participants had at least stage 3 CKD (eGFR <60 ml/min/1.73m^2^) and 433 (65.7%) had an eGFR >90 ml/min/1.73m^2^.

**Table 1.**
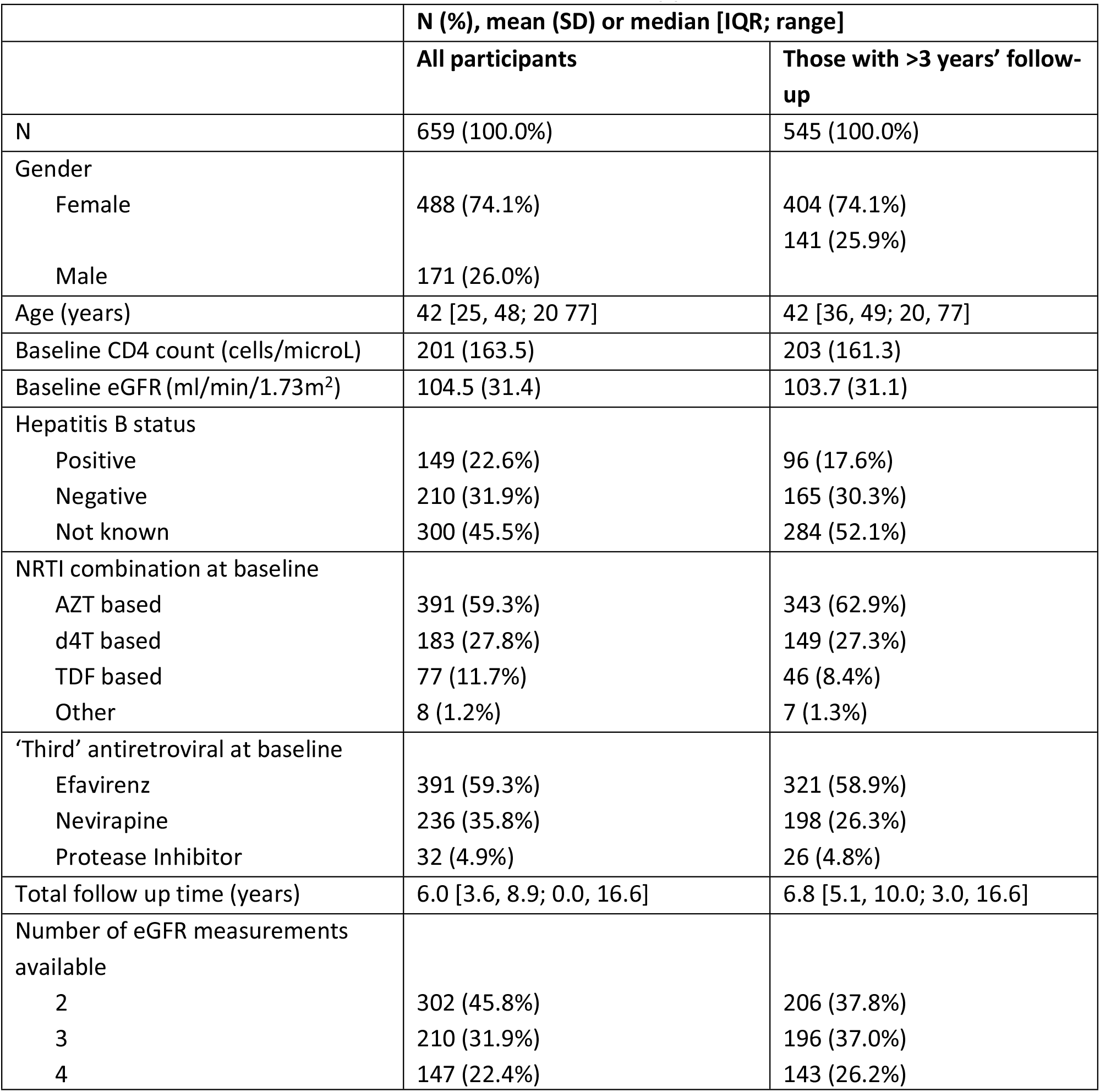
Patient characteristics at start of antiretroviral therapy.

|  | N (%), mean (SD) or median [IQR; range] |  |
| --- | --- | --- |
|  | All participants | Those with >3 years' follow-up |
| N | 659 (100.0%) | 545 (100.0%) |
| Gender |  |  |
| Female | 488 (74.1%) | 404 (74.1%) |
| Male | 171 (26.0%) | 141 (25.9%) |
| Age (years) | 42 [25, 48; 20 77] | 42 [36, 49; 20, 77] |
| Baseline CD4 count (cells/microL) | 201 (163.5) | 203 (161.3) |
| Baseline eGFR (ml/min/1.73m<sup>2</sup>) | 104.5 (31.4) | 103.7 (31.1) |
| Hepatitis B status |  |  |
| Positive | 149 (22.6%) | 96 (17.6%) |
| Negative | 210 (31.9%) | 165 (30.3%) |
| Not known | 300 (45.5%) | 284 (52.1%) |
| NRTI combination at baseline |  |  |
| AZT based | 391 (59.3%) | 343 (62.9%) |
| d4T based | 183 (27.8%) | 149 (27.3%) |
| TDF based | 77 (11.7%) | 46 (8.4%) |
| Other | 8 (1.2%) | 7 (1.3%) |
| 'Third' antiretroviral at baseline |  |  |
| Efavirenz | 391 (59.3%) | 321 (58.9%) |
| Nevirapine | 236 (35.8%) | 198 (26.3%) |
| Protease Inhibitor | 32 (4.9%) | 26 (4.8%) |
| Total follow up time (years) | 6.0 [3.6, 8.9; 0.0, 16.6] | 6.8 [5.1, 10.0; 3.0, 16.6] |
| Number of eGFR measurements available |  |  |
| 2 | 302 (45.8%) | 206 (37.8%) |
| 3 | 210 (31.9%) | 196 (37.0%) |
| 4 | 147 (22.4%) | 143 (26.2%) |

### Changes between first and last eGFR measurement – sub group followed for >3 years

Table 2 summarises the changes in eGFR measurements over time according to ART used and HBV status in the sub-group of 545 participants with at least three years follow-up. The mean increase over follow-up was 6.8 ml/min/1.73m^2^ (95% CI +4.0, +9.6 ml/min/1.73m^2^), equating to an annualised increase of 1.1 ml/min/1.73m^2^ per year (95% CI +0.6, +1.6; range -27.4, +29.6). There was considerable between-individual variability, with a standard deviation of 5.9 ml/min/1.73m^2^ per year. At final measurement, 17.5% of participants had reached at least stage 2 CKD (eGFR<90 ml/min/1.73m^2^) and 2.9% had reached at least CKD stage 3 (eGFR<60 ml/min/1.73m^2^).

**Table 2.**
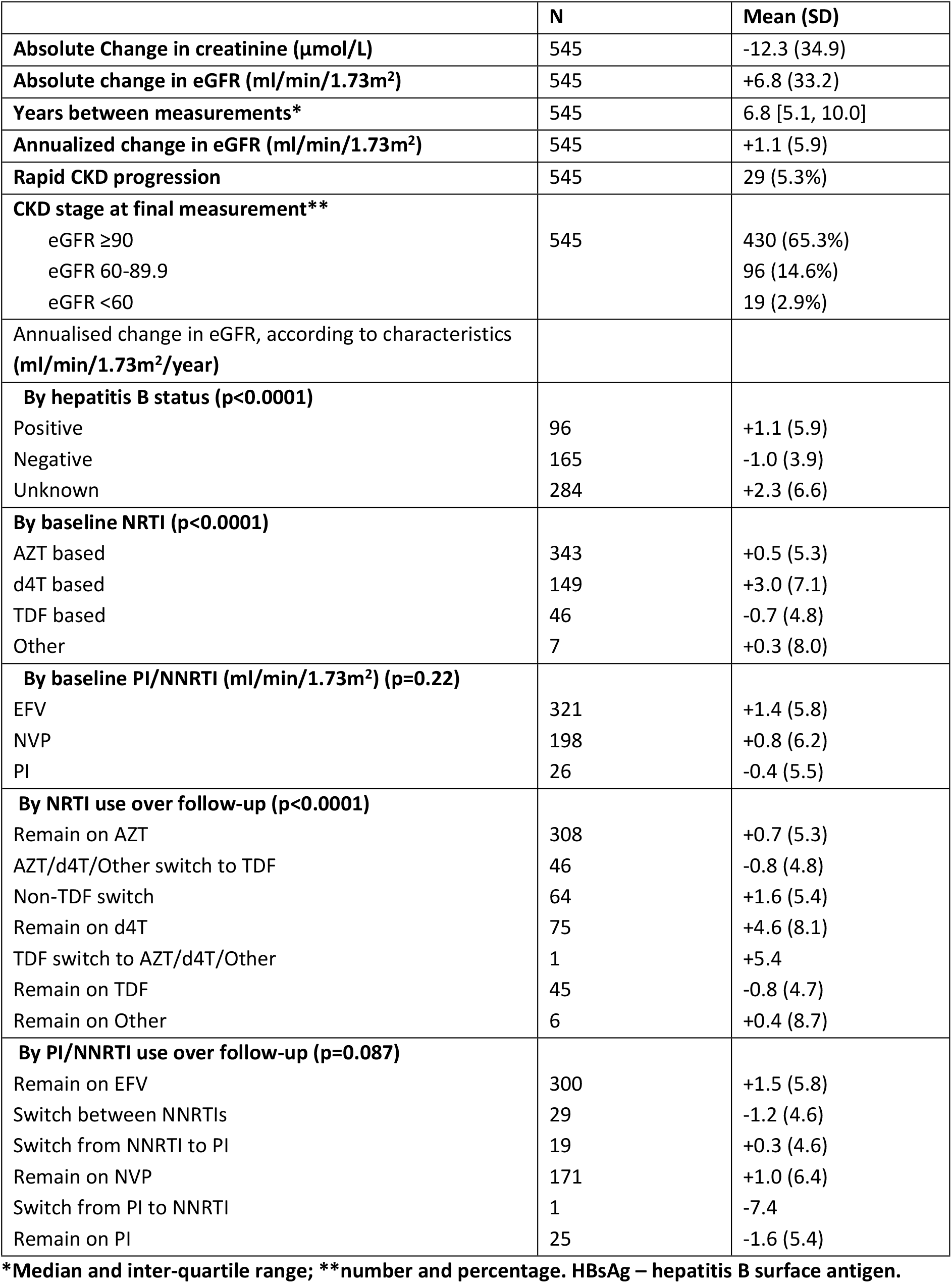
Evaluation of eGFR among those with at least 3 years’ follow-up, according to ART regimen and HBsAg status.

|  | **N** | **Mean (SD)** |
| --- | --- | --- |
| **Absolute Change in creatinine ($\mu\text{mol/L}$)** | 545 | -12.3 (34.9) |
| **Absolute change in eGFR ($\text{ml/min/1.73m}^2$)** | 545 | +6.8 (33.2) |
| **Years between measurements\*** | 545 | 6.8 [5.1, 10.0] |
| **Annualized change in eGFR ($\text{ml/min/1.73m}^2$)** | 545 | +1.1 (5.9) |
| **Rapid CKD progression** | 545 | 29 (5.3%) |
| **CKD stage at final measurement\*\*** |  |  |
| eGFR $\geq 90$ | 545 | 430 (65.3%) |
| eGFR 60-89.9 |  | 96 (14.6%) |
| eGFR $< 60$ | | 19 (2.9%) |
| **Annualised change in eGFR, according to characteristics ($\text{ml/min/1.73m}^2/\text{year}$)** |  |  |
| **By hepatitis B status ($p < 0.0001$)** |  |  |
| Positive | 96 | +1.1 (5.9) |
| Negative | 165 | -1.0 (3.9) |
| Unknown | 284 | +2.3 (6.6) |
| **By baseline NRTI ($p < 0.0001$)** |  |  |
| AZT based | 343 | +0.5 (5.3) |
| d4T based | 149 | +3.0 (7.1) |
| TDF based | 46 | -0.7 (4.8) |
| Other | 7 | +0.3 (8.0) |
| **By baseline PI/NNRTI ($\text{ml/min/1.73m}^2$) ($p = 0.22$)** |  |  |
| EFV | 321 | +1.4 (5.8) |
| NVP | 198 | +0.8 (6.2) |
| PI | 26 | -0.4 (5.5) |
| **By NRTI use over follow-up ($p < 0.0001$)** |  |  |
| Remain on AZT | 308 | +0.7 (5.3) |
| AZT/d4T/Other switch to TDF | 46 | -0.8 (4.8) |
| Non-TDF switch | 64 | +1.6 (5.4) |
| Remain on d4T | 75 | +4.6 (8.1) |
| TDF switch to AZT/d4T/Other | 1 | +5.4 |
| Remain on TDF | 45 | -0.8 (4.7) |
| Remain on Other | 6 | +0.4 (8.7) |
| **By PI/NNRTI use over follow-up ($p = 0.087$)** |  |  |
| Remain on EFV | 300 | +1.5 (5.8) |
| Switch between NNRTIs | 29 | -1.2 (4.6) |
| Switch from NNRTI to PI | 19 | +0.3 (4.6) |
| Remain on NVP | 171 | +1.0 (6.4) |
| Switch from PI to NNRTI | 1 | -7.4 |
| Remain on PI | 25 | -1.6 (5.4) |

\*Median and inter-quartile range; \*\*number and percentage. HBsAg – hepatitis B surface antigen.

Table 2 also shows the mean annualised eGFR change according to hepatitis B status and ART use. There was strong evidence of a difference in eGFR changes over follow-up according to hepatitis B status; although this association was mostly driven by an increase among those of unknown status, there was also evidence of a recovery in eGFR in HBsAg positive individuals compared to HBsAg negative individuals. There was also evidence of greater increases in those receiving d4T-based NRTI backbones. There was no evidence of an association with specific NNRTI or PI use.

### Prevalence of rapid eGFR progression

Table 3 provides information on the proportion of participants in each subgroup who demonstrated rapid eGFR deterioration in the sub-group with >3 years’ follow-up. In total, just 5.3% (29/545) experienced an annualised decline in eGFR of >5 ml/min/1.73m^2^/year, as well as a final eGFR value <90 ml/min/1.73m^2^. Although numbers were small, there was some evidence of an association between rapid progression and PI/NNRTI use over follow-up: the proportions of participants developing rapid eGFR decline ranged from 4% (in those continuing on a PI) to 100% (in those switching from a PI to a NNRTI), albeit with a sample size of 1 in the latter case (p=0.069).

**Table 3.** Factors associated with eGFR change per year: results from multi-level model incorporating all eGFR measurements recorded over a period up to 15 years.

|  |  | Univariable |  |  | Multivariable |  |  |
| --- | --- | --- | --- | --- | --- | --- | --- |
|  |  | Beta | 95% CI | P | Beta | 95% CI | P |
| Gender (vs male) | Female | -0.47 | -1.10, 0.17 | 0.15 | -0.31 | -0.96, 0.33 | 0.35 |
| Baseline age | Per 10 years | -0.17 | -0.46, 0.12 | 0.25 | 0.13 | -0.19, 0.44 | 0.43 |
| Current NRTI (vs AZT) | d4T | 1.69 | 0.73, 2.66 | <0.0001 | 1.21 | 0.25, 2.16 | 0.008 |
|  | TDF | -1.60 | -2.39, -0.80 |  | -1.08 | -1.92, -0.24 |  |
|  | Other | 0.69 | -1.54, 2.92 |  | 1.53 | -0.92, 3.99 |  |
| Current Third drug (vs EFV) | NVP | -0.77 | -1.38, -0.16 | 0.0009 | -0.78 | -1.39, -0.17 | 0.0029 |
|  | PI | -1.73 | -2.76, -0.70 |  | -1.55 | -2.68, -0.41 |  |
| Hepatitis B (vs Negative) | Positive | 1.51 | 0.56, 2.46 | <0.0001 | 1.25 | 0.29, 2.20 | <0.0001 |
|  | Unknown | 1.65 | 1.07, 2.24 |  | 1.38 | 0.74, 2.01 |  |

### Factors associated with long-term changes in eGFR

Multi-level models considering all 659 participants identified that treatment with TDF was associated with the poorest eGFR outcomes of all NRTIs (Table 4), with a statistically significant eGFR greater decline per year of –1.08ml/min/1.73 m^2^ (CI –1.92, -0.24) compared with zidovudine (AZT). TDF was also the only drug which was associated with a net negative eGFR change irrespective of whether it was commenced immediately after diagnosis of HIV or switched onto from a different NRTI (Table 2). In contrast, stavudine (d4T) was associated with relatively favourable eGFR outcomes, with an average change per year that was +1.21ml/min/1.73 m^2^ (95% CI +0.25, +2.16) greater compared with AZT. The analysis found no significant difference in relative eGFR change over time between AZT and pooled results from the remaining less common NRTIs (difference –1.53 ml/min/1.73 m^2^, 95% CIs –0.92, 3.99). There was also evidence that NVP and PI use was associated with lower rates of change per year compared to EFV and that positive and unknown HBsAg status was associated with higher eGFR changes per year, compared to HBsAg negative status.

### Sensitivity analysis

Sensitivity analysis considering the multi-level model restricted to the sub-group with longer follow up found consistent results with the main findings (see Table 5).

## Discussion

Our results support previous findings highlighting that where eGFRs decline, TDF and protease inhibitors have associations with greater eGFR declines compared with other antiretroviral drugs^27,34,35^. Nevirapine also showed significantly greater associations with eGFR decline compared to efavirenz and there were significant differences in proportions of participants who experienced rapid eGFR decline. Our findings provide evidence that in most cases significant eGFR decline is infrequent during the first years of treatment.

An interesting finding of this study was a large initial increase in eGFR across the cohort following ART followed by a modest decline across subsequent eGFR measurements (figure 1); similar results have been noted previously from studies on similar populations^42,43^. Our findings most likely reflect a combination of factors relating to studying a population with considerably higher rates of advanced HIV infection (leading to more frequent HIV-associated nephropathies) at diagnosis compared with HIC cohorts. Firstly, survivor bias is likely to have caused an artificial ‘increase’ in mean eGFR if those with the most advanced disease passed away shortly following initial measurement, leaving those who are able to have further eGFR measurements to generate a higher mean value by default. A further factor might be due to reversable eGFR function in successfully treated participants who were initially acutely unwell. Once ART was commenced and they clinically improved, renal function may have returned to a higher baseline level, overshadowing other contributors towards eGFR decline. Further research is needed to explore this effect more closely, and future studies on ARTs in LMICs should take this into account in their methodology and analysis. It is important to note that in spite of the overall increase in eGFR across our participants, TDF was the only drug noted to still demonstrate negative associations with eGFR decline irrespective whether it was started at baseline or if switched to at a later date. This strongly supports the case for more rapidly switching ART regimens to include tenofovir alafenamide (TAF), and integrase inhibitors, which are now more widely available in Ghana and sub-Saharan Africa.

**Figure 1.**
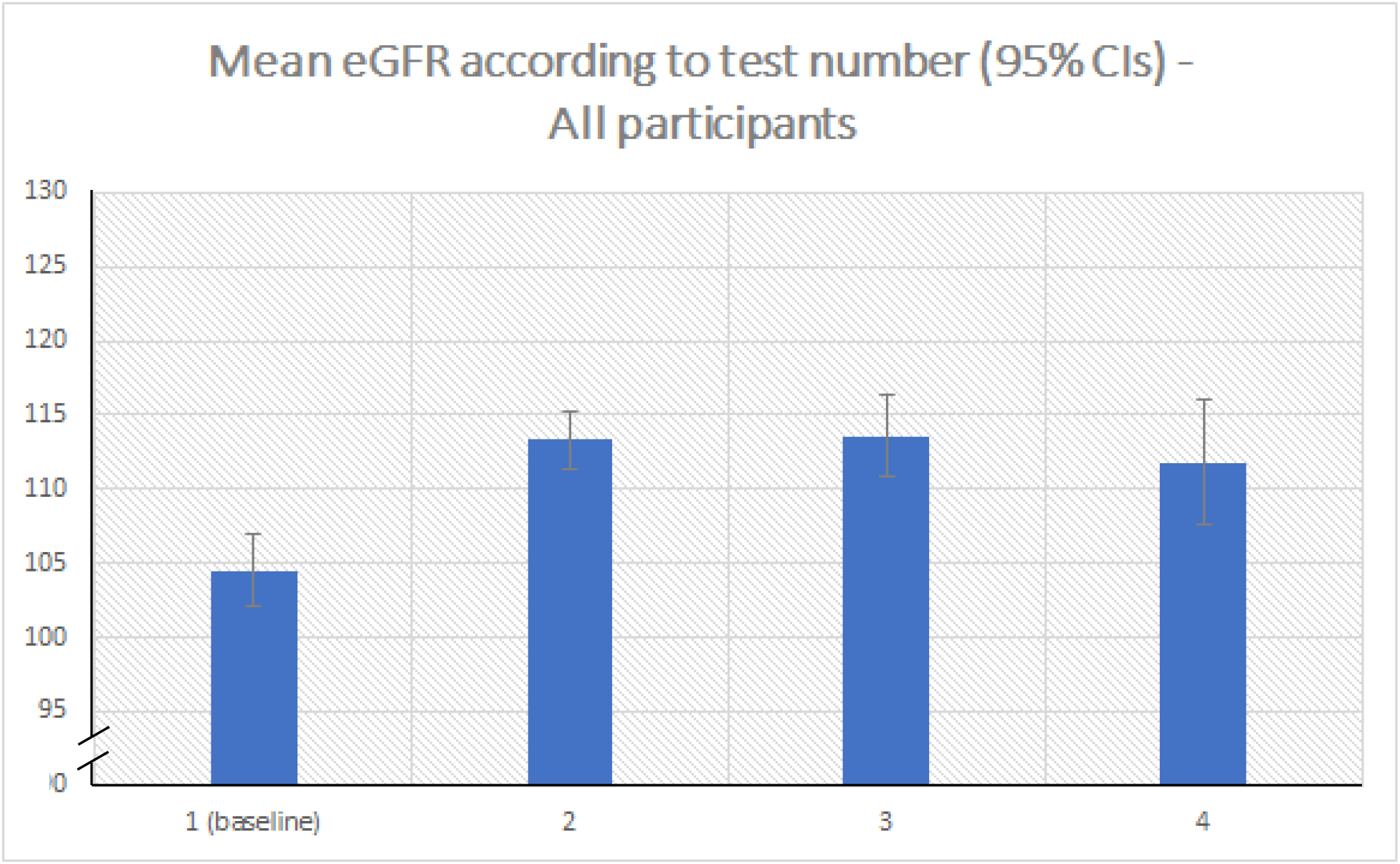
Mean eGFR scores in all participants for baseline, second, third and fourth blood tests, demonstrating an initial sharp mean increase in eGFR post-treatment followed by a gradual mean decline. NB: a bar graph was selected because time between measures was not accounted for in this graph.

The finding of a positive effect HBsAg co-infection on eGFR outcomes was unexpected based on our initial hypotheses and is not consistent with evidence from the literature assessing HBV’s association with CKD^44^. Post-hoc analysis of our results highlighted a mean eGFR increase of almost 10 ml/min/1.73m^2^ between first and second blood tests across HBV positive participants but no increase in HBV negative participants (supplementary figure 1). We believe sampling bias may have had an influence on these results. HBV testing has not historically been routine practice in HIV management in Ghana; a main driving factor for tests being requested in PWH (outside of research studies) was when participants had elevated transaminase enzymes, a proxy marker of underlying liver pathology. As a result, patients with elevated transaminase enzymes but a negative HBsAg test may have had alternative serious comorbidities contributing to less significant improvement in eGFR post-treatment or more significant declines thereafter. Another factor underlying this finding is indication bias; PWH found to be HBsAg positive were much more likely to be prescribed tenofovir, a drug with well-documented associations with eGFR decline in the literature^32-34^.

There were several limitations to our study. Our study cohort was relatively young, with less than a quarter of participants being above 50 years old. There is some evidence that renal function may decline at faster rates as people age^45^, which suggests our measures of eGFR decline may not reflect real-world rates in older PWH. A younger cohort is also likely to underestimate incidence of CKD development as participants’ baseline kidney functions are more frequently well above CKD-relevant eGFR thresholds.

Despite being a major element of the KDIGO CKD scoring criteria, proteinuria was not included as an outcome variable in our study. This was because testing for proteinuria was not routine in the clinic so not all patients in the convenience and research cohorts had proteinuria measurements. While evidence exists noting associations of proteinuria with use of ART regimens such as tenofovir^38,46^, studies of adequate duration are still sparse in LMICs and more long-term research is required. The need for a greater focus towards prevention and early detection of CKD in low- and middle-income countries has been emphasized in global health literature^47,48^, in part because treatment for ESRD is highly specialized^49^ and costly for healthcare systems already challenged with scarcer resources than HICs. This is particularly important since other comorbidities contributing to CKD are increasingly seen in this population^50,51^.

Our research was conducted in an LMIC region (with less accessible healthcare than HICs) so findings were reliant on a relatively low number of creatinine results per patient. In addition, it is probable that fewer eGFR tests would have been taken as part of routine testing, with a relatively higher percentage of tests being taken when a patient is unwell, adding noise to study findings. Other possible influencers of eGFR change, such as adherence to ART, or diseases like diabetes hypertension and TB, were not assessed in this study and may have led to confounding.

We included participants from a range of different study cohorts including HEPIK^27^, as well as an additional convenience sample specific to this study. This means that it did not have all the characteristics of a uniform cohort study. In addition, our ability to follow-up participants for additional results was limited so we could not observe important outcomes such as mortality rates, clinical events or other reasons for loss to follow-up. These data would have been important in generating a clearer picture of the real-world effects of ART and HBV status on renal function. We included participants through a variety of studies that did not always share identical inclusion criteria. This may have led to selection bias, most likely through over-sampling HBV-positive participants into our pooled cohort.

Lastly, the cohort nature of the study leaves it vulnerable to the effects of confounding, given relatively limited data were available on other comorbidities, co-medications or other factors that can affect eGFR.

## Conclusions

This study is one of the first long-term analyses of the effects of ART and HBV status on renal function in a sub-Saharan African population. It supports evidence that TDF and protease inhibitors have associations with worsening renal function over time compared with other antiretroviral drugs. However, our study highlights the importance of future LMIC-focused studies accounting for significant effects of survivorship bias and/or initial improvements in eGFR affecting findings when establishing their methodology.

Establishing the most significant contributors towards renal disease is an essential means to be able to minimise its overall health burden. Small discrepancies in annual eGFR decline may accumulate over time leading to considerably higher rates of CKD in older populations of PLH. The prevalence of non-communicable diseases such as diabetes and hypertension are increasing in LMICs which will place further burden on the renal health of PWH. Long-term prospective studies encompassing such factors are essential to closely assess contributors to renal function decline in LMIC populations.

## Data Availability

Data can be accessed through emailing corresponding author and can be made publicly accessible if necessary

## Acknowledgements

We are grateful to the nurses and patients at KATH HIV clinic for assistance with this study. We would also like to acknowledge the following for assistance with data and sample collection: David Stewart, Jess Lundgren, Ryhan Hussein and Shelley Burton.

## Authors’ contributions

DRC designed the study and assisted with manuscript preparation, sample analysis, data collection and data interpretation. FB was the primary writer of the manuscript and assisted with sample analysis, data collection and data interpretation. CS assisted with study design, analysed the data, assisted with data interpretation and critically evaluated the manuscript. DO, GV, OP and YH assisted with sample analysis and data collection and critically evaluated the manuscript. RP, AMG and SS assisted with study design and critically evaluated the manuscript.

## List of Abbreviations

HIV: Human immunodeficiency virus
PWH: People living with HIV
HIVAN: HIV-associated nephropathy
ART: Antiretroviral therapy
ESRD: End stage renal disease
HBV: Hepatitis B virus

## Supplementary Tables

**Supplementary table 1 -.** Frequency of rapid progression (RP) during study period among those with >3 years’ follow-up, according to ART regimen and HBV status.

|  | n/N (%) | p-value |
| --- | --- | --- |
| Total | 29/545 |  |
| **According to hepatitis B status** |  |  |
| Positive | 6/96 (6.3%) | 0.89 |
| Negative | 9/165 (5.5%) |  |
| Unknown | 14/284 (4.9%) |  |
| **According to baseline NRTI** |  |  |
| AZT based | 21/343 (6.1%) | 0.55 |
| d4T based | 5/149 (3.4%) |  |
| TDF based | 3/46 (6.5%) |  |
| Other | 0/7 (0.0%) |  |
| **According to baseline PI/NNRTI** |  |  |
| EFV | 16/321 (5.0%) | 0.35 |
| NVP | 10/198 (5.1%) |  |
| PI | 3/26 (11.5%) |  |
| **According to NRTI over follow-up** |  |  |
| Remain on AZT | 16/308 (5.2%) | 0.61 |
| AZT/d4T/Other switch to TDF | 5/46 (10.9%) |  |
| Non-TDF switch | 3/64 (4.7%) |  |
| Remain on d4T | 2/75 (2.7%) |  |
| TDF switch to AZT/d4T/Other | 0/1 (0.0%) |  |
| Remain on TDF | 3/45 (6.7%) |  |
| Remain on Other | 0/6 (0.0%) |  |
| **According to PI/NNRTI over follow-up** |  |  |
| Remain on EFV | 14/300 (4.7%) | 0.069 |
| Switch between NNRTIs | 3/29 (10.3%) |  |
| Switch from NNRTI to PI | 1/19 (5.3%) |  |
| Remain on NVP | 8/171 (4.7%) |  |
| Switch from PI to NNRTI | 1/1 (100.0%) |  |
| Remain on PI | 2/25 (4.0%) |  |

**Supplementary Table 2.** Multivariable sensitivity analysis where only participants with follow-up period spanning >3 years were included in analysis.

|  |  | Univariable |  |  | Multivariable |  |  |
| --- | --- | --- | --- | --- | --- | --- | --- |
|  |  | Beta | 95% CI | P | Beta | 95% CI | P |
| Gender (vs male) | Female | -0.38 | -1.04, 0.27 | 0.25 | -0.32 | -0.98, 0.34 | 0.34 |
| Baseline age | Per 10 years | -0.18 | -0.47, 0.12 | 0.23 | 0.11 | -0.21, 0.43 | 0.51 |
| Current NRTI (vs AZT) | d4T | 1.67 | 0.70, 2.65 | <0.0001 | 1.20 | 0.23, 2.16 | 0.004 |
|  | TDF | -1.46 | -2.31, -0.61 |  | -1.00 | -1.90, -0.11 |  |
|  | Other | 0.22 | -2.04, 2.49 |  | 1.06 | -0.44, 3.56 |  |
| Current Third drug (vs EFV) | NVP | -0.77 | -1.39, -0.15 | 0.0005 | -0.73 | -1.35, -0.10 | 0.005 |
|  | PI | -1.90 | -2.95, -0.84 |  | -1.60 | -2.78, -0.41 |  |
| Hepatitis B (vs Negative) | Positive | 1.27 | 0.29, 2.26 | <0.0001 | 1.07 | 0.07, 2.06 | 0.0003 |
|  | Unknown | 1.48 | 0.89, 2.08 |  | 1.32 | 0.66, 1.96 |  |

**Supplementary Figure 1 -.**
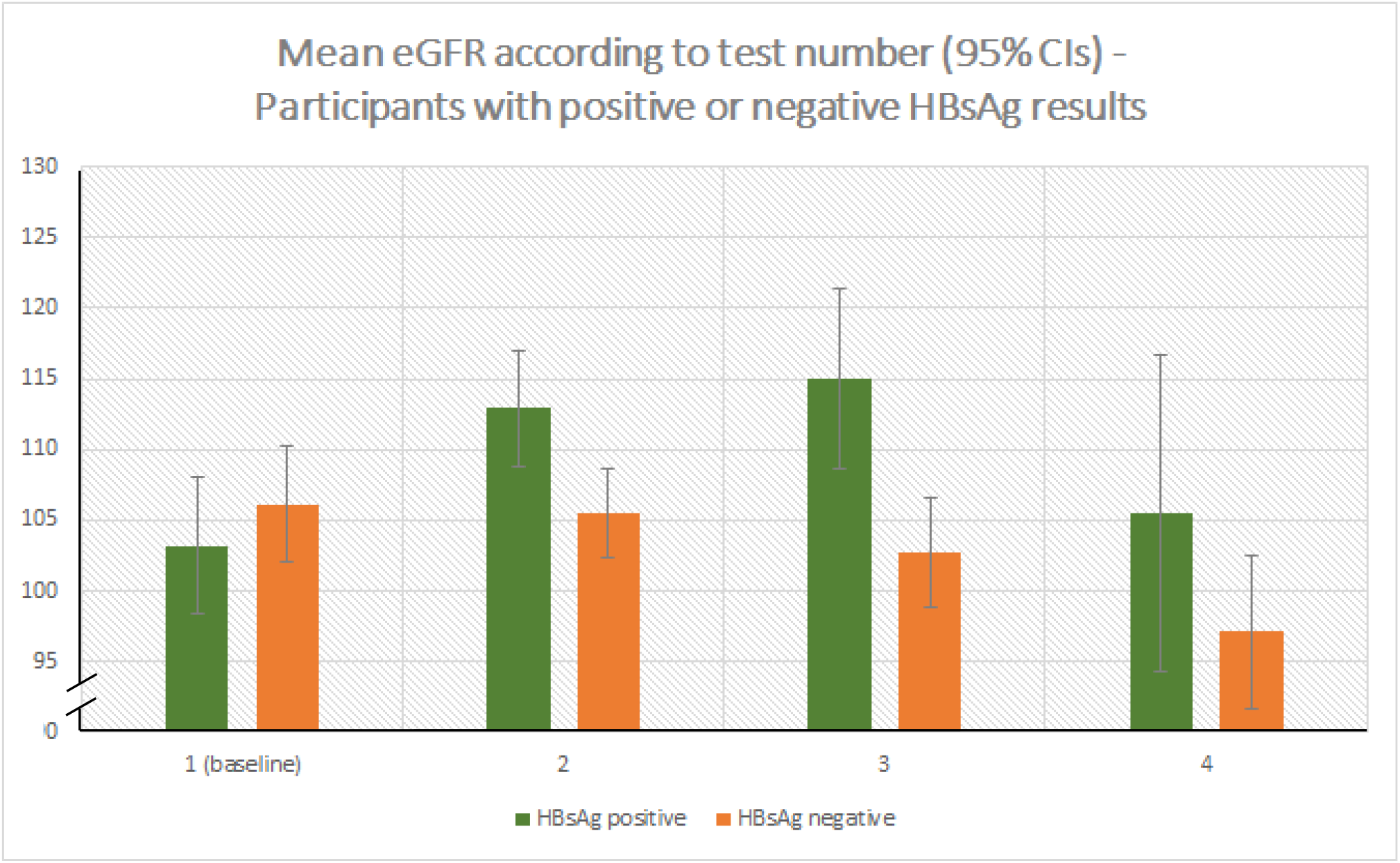
Post-hoc analysis of mean eGFR scores in HBsAg positive and negative participants for baseline, second, third and fourth bloods tests, highlighting that HBsAg positive participants demonstrated a larger increase in eGFR following ART than the HBsAg negative subgroup.

